# Association between social media use and self-reported symptoms of depression in US adults

**DOI:** 10.1101/2021.10.26.21265528

**Authors:** Roy H. Perlis, Jon Green, Matthew Simonson, Katherine Ognyanova, Mauricio Santillana, Jennifer Lin, Alexi Quintana, Hanyu Chwe, James Druckman, David Lazer, Matthew A. Baum, John Della Volpe

## Abstract

**Importance:** Some studies suggest that social media use is associated with risk for depression, particularly among children and young adults.

**Objective:** To characterize the association between self-reported use of individual social media platforms, and worsening of depressive symptoms, among adults.

**Design:** We included data from 13 waves of a non-probability internet survey conducted between May 2020 and May 2021 among individuals age 18 and older in the United States. We applied logistic regression with 5 or more point increase in PHQ-9 as outcome, and sociodemographic features, baseline PHQ-9, and use of each social media platform as independent variables, without reweighting.

**Participants:** Survey respondents 18 and older.

**Main Outcome and Measure:** PHQ-9 worsening by 5 points or more.

**Results:** A total of 5395/8045 (67.1%) individuals with a PHQ-9<5 on initial survey completed a second PHQ-9. These respondents had mean age 55.82 years (SD 15.17); 3546 (65.7%) reported female gender, the remainder male; 329 (6.1%) were Asian, 570 (10.6%) Black, 256 (4.7%) Hispanic, 4118 (76.3%) White, and 122 (2.3%) Native American or Alaska Native, Pacific Islander or Native Hawaiian, or Other. Among them, 482 (8.9%) reported 5 point or greater worsening at second survey. In fully-adjusted models for increase in symptoms, largest odds ratio associated with social media use was observed for Snapchat (aOR 1.53, 95% CI 1.19-1.96), Facebook (aOR 1.42, 1.10-1.81), and TikTok (aOR 1.39, 95% CI 1.03-1.87).

**Conclusions and Relevance:** Among survey respondents who did not report depressive symptoms initially, social media use was associated with greater likelihood of subsequent increase in depressive symptoms after adjustment for sociodemographic features and news sources. These data cannot elucidate the nature of this relationship, but suggest the need for further study to understand the impact of social media use.

**Trial Registration:** NA

## Introduction

Social media use has been associated with diminished well-being and greater levels of anxiety and depression, predominantly in cross-sectional studies among adolescents^1^ or young adults^2^, although concern has been raised for reporting bias^3^, if individuals with greater depressive symptoms overreport social media use. A small number of short-term longitudinal studies provide further support for this association^4,5^ – for example, among 82 young adults sampled 2 weeks apart.

These results have two notable gaps. First, is the observed cross-sectional relationship also apparent in longitudinal studies, as suggested by a randomized trial of Facebook discontinuation^6^? Second, does this risk apply to older consumers of social media? To investigate these questions, we utilized data from multiple waves of an ongoing 50-state US survey.

## Method

Data were drawn from 13 waves of a non-probability internet survey deployed via a multi-panel commercial vendor approximately every month between May 2020 and May 2021 in individuals 18 and older. The study was approved by the Institutional Review Board of Harvard University, and participants provided informed consent.

The sampling strategy incorporated quotas for gender, age, and race/ethnicity within each state; attention checks and open-ended answers were used to filter out unreliable respondents. Race/ethnicity was self-reported from 5 Census categories to confirm representativeness of the US population. Participants also completed the 9-item Patient Health Questionnaire (PHQ-9)^7^. They were asked, “Do you ever use any of the following social media sites or apps?”; we focused a priori on Facebook, Instagram, LinkedIn, Pinterest, TikTok, Twitter, Snapchat, and YouTube. They were further asked to identify any sources of COVID-19 related news in the past 24 hours (here, cable or network television, or web news site), which we used as a proxy for news sources more generally, number of social supports available “to talk to if you had a problem, felt sad or depressed”, and face-to-face meetings with non-household members in the prior 24 hours. (As some survey modules were randomly assigned to a subset of participants in each wave in order to diminish overall survey length, n=1646 returning participants completed news source, and n=4808 completed social supports questions; Table 1).

**Table 1.**
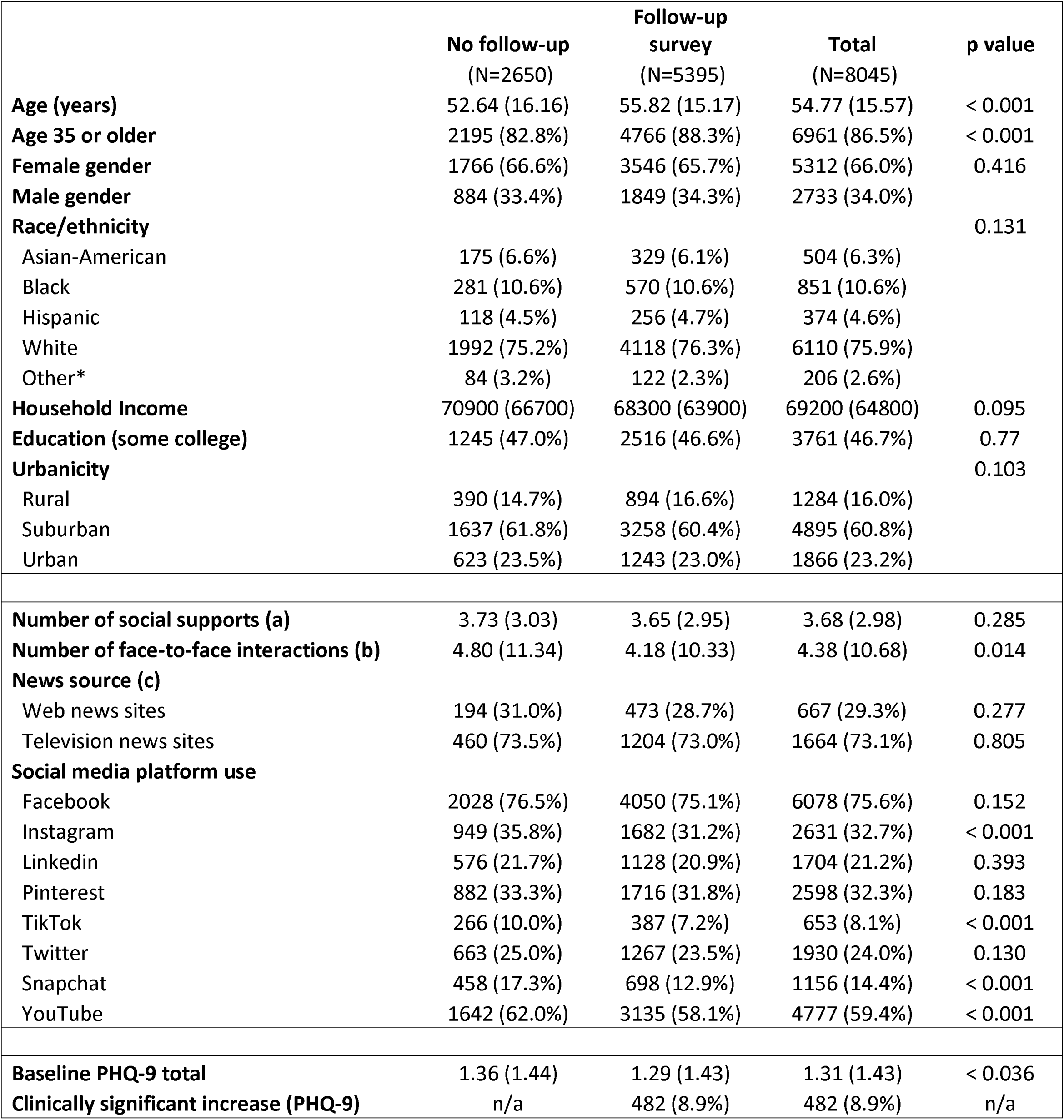
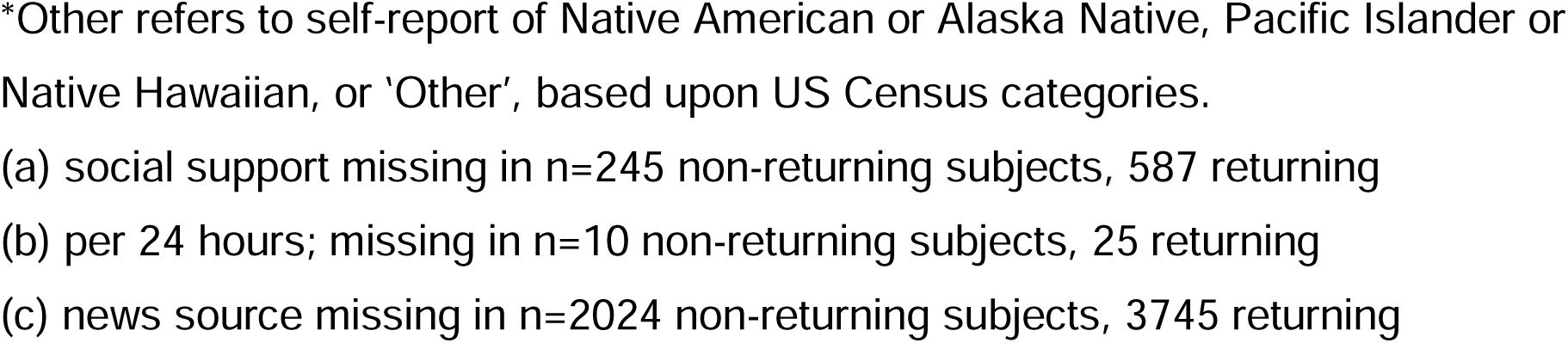
Comparison of survey respondents who did, or did not, complete PHQ-9 at a subsequent wave after initial PHQ-9

|  | **No follow-up**<br>(N=2650) | **Follow-up survey**<br>(N=5395) | **Total**<br>(N=8045) | **p value** |
| --- | --- | --- | --- | --- |
| **Age (years)** | 52.64 (16.16) | 55.82 (15.17) | 54.77 (15.57) | < 0.001 |
| **Age 35 or older** | 2195 (82.8%) | 4766 (88.3%) | 6961 (86.5%) | < 0.001 |
| **Female gender** | 1766 (66.6%) | 3546 (65.7%) | 5312 (66.0%) | 0.416 |
| **Male gender** | 884 (33.4%) | 1849 (34.3%) | 2733 (34.0%) |  |
| **Race/ethnicity** |  |  |  | 0.131 |
| Asian-American | 175 (6.6%) | 329 (6.1%) | 504 (6.3%) |  |
| Black | 281 (10.6%) | 570 (10.6%) | 851 (10.6%) |  |
| Hispanic | 118 (4.5%) | 256 (4.7%) | 374 (4.6%) |  |
| White | 1992 (75.2%) | 4118 (76.3%) | 6110 (75.9%) |  |
| Other* | 84 (3.2%) | 122 (2.3%) | 206 (2.6%) |  |
| **Household Income** | 70900 (66700) | 68300 (63900) | 69200 (64800) | 0.095 |
| **Education (some college)** | 1245 (47.0%) | 2516 (46.6%) | 3761 (46.7%) | 0.77 |
| **Urbanicity** |  |  |  | 0.103 |
| Rural | 390 (14.7%) | 894 (16.6%) | 1284 (16.0%) |  |
| Suburban | 1637 (61.8%) | 3258 (60.4%) | 4895 (60.8%) |  |
| Urban | 623 (23.5%) | 1243 (23.0%) | 1866 (23.2%) |  |
| **Number of social supports (a)** | 3.73 (3.03) | 3.65 (2.95) | 3.68 (2.98) | 0.285 |
| **Number of face-to-face interactions (b)** | 4.80 (11.34) | 4.18 (10.33) | 4.38 (10.68) | 0.014 |
| **News source (c)** |  |  |  |  |
| Web news sites | 194 (31.0%) | 473 (28.7%) | 667 (29.3%) | 0.277 |
| Television news sites | 460 (73.5%) | 1204 (73.0%) | 1664 (73.1%) | 0.805 |
| **Social media platform use** |  |  |  |  |
| Facebook | 2028 (76.5%) | 4050 (75.1%) | 6078 (75.6%) | 0.152 |
| Instagram | 949 (35.8%) | 1682 (31.2%) | 2631 (32.7%) | < 0.001 |
| Linkedin | 576 (21.7%) | 1128 (20.9%) | 1704 (21.2%) | 0.393 |
| Pinterest | 882 (33.3%) | 1716 (31.8%) | 2598 (32.3%) | 0.183 |
| TikTok | 266 (10.0%) | 387 (7.2%) | 653 (8.1%) | < 0.001 |
| Twitter | 663 (25.0%) | 1267 (23.5%) | 1930 (24.0%) | 0.130 |
| Snapchat | 458 (17.3%) | 698 (12.9%) | 1156 (14.4%) | < 0.001 |
| YouTube | 1642 (62.0%) | 3135 (58.1%) | 4777 (59.4%) | < 0.001 |
| **Baseline PHQ-9 total** | 1.36 (1.44) | 1.29 (1.43) | 1.31 (1.43) | < 0.036 |
| **Clinically significant increase (PHQ-9)** | n/a | 482 (8.9%) | 482 (8.9%) | n/a |

\*Other refers to self-report of Native American or Alaska Native, Pacific Islander or Native Hawaiian, or 'Other', based upon US Census categories.
(a) social support missing in n=245 non-returning subjects, 587 returning
(b) per 24 hours; missing in n=10 non-returning subjects, 25 returning
(c) news source missing in n=2024 non-returning subjects, 3745 returning

The study cohort included participants who completed the PHQ-9 in at least two survey waves with PHQ-9 total score<5 (less than mild depression) at the index survey. We applied logistic regression with 5 or more point increase in PHQ-9, the threshold for clinical significance^7^, as outcome, and sociodemographic features, baseline PHQ-9, and use of each social media platform as independent variables using *glm* (R 3.6), without reweighting. Reporting followed AAPOR guidelines: https://www.aapor.org/Standards-Ethics/AAPOR-Code-of-Ethics/Survey-Disclosure-Checklist.aspx.

## Results

Overall, 5395/8045 (67.1%) individuals with PHQ-9<5 on initial survey completed a subsequent PHQ-9 (Table 1). These respondents had mean age 55.82 years (SD 15.17); 3546 (65.7%) reported female gender, the remainder male; 329 (6.1%) were Asian, 570 (10.6%) Black, 256 (4.7%) Hispanic, 4118 (76.3%) White, and 122 (2.3%) Native American or Alaska Native, Pacific Islander or Native Hawaiian, or “Other”. Mean PHQ-9 score at initial survey for this group of subsequent responders was 1.29 (SD 1.43). At first follow-up survey, 482 (8.9%) experienced an increase in PHQ-9 of 5 points or greater.

In adjusted regression models, Snapchat, Facebook, and TikTok use at first survey were significantly associated with greater risk of increase in self-reported depressive symptoms (Table 2, left), with adjusted odds ratios of 1.53 (95% CI 1.19-1.96), 1.42, (95% CI 1.10-1.81), and 1.39 (95% CI 1.03-1.87), respectively. Incorporating terms for television or internet news in the past 24 hours, number of social supports, and number of daily face-to-face interactions on the initial survey did not meaningfully change these associations (Table 2, right) with the notable exception of Snapchat, where adjusted odds ratio (aOR) was diminished from 1.53 (95% CI 1.19-1.96) to 1.12 (95% CI 0.70-1.81) with the inclusion of terms for news source.

**Table 2.** Effect sizes for social media platform, without (left) or with (right) adjustment for social support, social interactions, and news source

| Platform | Full cohort |  |  |  | With News Sources^ |  |  | With Social Support |  |  | With Social Interactions |  |
| --- | --- | --- | --- | --- | --- | --- | --- | --- | --- | --- | --- | --- |
|  | OR | [95% CI ] | P value |  | OR | [95% CI ] |  | OR | [95% CI ] |  | OR | [95% CI ] |
| Facebook | 1.42 | 1.10 1.81 | .007 |  | 1.38 | 0.88 2.19 |  | 1.51 | 1.16 1.97 |  | 1.40 | 1.09 1.79 |
| Instagram | 1.18 | 0.95 1.46 | 0.13 |  |  |  |  |  |  |  |  |  |
| Linkedin | 1.03 | 0.80 1.33 | 0.24 |  |  |  |  |  |  |  |  |  |
| Pinterest | 0.94 | 0.76 1.16 | 0.57 |  |  |  |  |  |  |  |  |  |
| TikTok | 1.39 | 1.03 1.87 | 0.03 |  | 1.40 | 0.78 2.52 |  | 1.41 | 1.03 1.94 |  | 1.38 | 1.02 1.86 |
| Twitter | 1.05 | 0.84 1.31 | 0.41 |  |  |  |  |  |  |  |  |  |
| Snapchat | 1.53 | 1.19 1.96 | <.001 |  | 1.12 | 0.70 1.81 |  | 1.61 | 1.23 2.10 |  | 1.52 | 1.19 1.96 |
| YouTube | 1.16 | 0.93 1.44 | 0.18 |  |  |  |  |  |  |  |  |  |

^ news sources only available for n=1646 respondents

In logistic regression models for increase in depressive symptoms, we further identified significant interactions of platform use with age group for Facebook, TikTok, and Snapchat (Table 3). For TikTok and Snapchat, greater magnitude of association with increase in depressive symptoms was identified among those age 35 or older compared to those younger than age 35. For Facebook, the opposite pattern was observed, with higher effect sizes for utilization observed among those younger than age 35 (aOR 2.60 (95% CI 1.46-1.62, versus aOR 1.12, 95% CI 0.85-1.48).

**Table 3.** Logistic regression models examining association between social media use at initial survey and clinically significant (5-point or greater) increase in PHQ-9

| Platform | Full cohort |  |  | age interact P | age 35+ |  |  |  | age< 35 |  |  |
| --- | --- | --- | --- | --- | --- | --- | --- | --- | --- | --- | --- |
|  | OR | [95% CI ] |  | value | OR | [95% CI ] |  |  | OR | [95% CI ] |  |
| Facebook | 1.42 | 1.10 | 1.81 | 0.04 | 1.12 | 0.85 | 1.48 |  | 2.60 | 1.46 | 4.62 |
| Instagram | 1.18 | 0.95 | 1.46 | 0.17 |  |  |  |  |  |  |  |
| Linkedin | 1.03 | 0.80 | 1.33 | 0.08 |  |  |  |  |  |  |  |
| Pinterest | 0.94 | 0.76 | 1.16 | 0.20 |  |  |  |  |  |  |  |
| TikTok | 1.39 | 1.03 | 1.87 | 0.01 | 1.67 | 1.14 | 2.45 |  | 1.36 | 0.86 | 2.16 |
| Twitter | 1.05 | 0.84 | 1.31 | 0.08 |  |  |  |  |  |  |  |
| Snapchat | 1.53 | 1.19 | 1.96 | <0.001 | 1.96 | 1.44 | 2.65 |  | 1.17 | 0.78 | 1.77 |
| YouTube | 1.16 | 0.93 | 1.44 | <0.001 | 1.31 | 1.03 | 1.67 |  | 0.68 | 0.41 | 1.11 |

OR, odds ratio; CI, confidence interval. Cohort includes all participants with PHQ-9 less than 5 at first survey who responded to a subsequent survey. Age-stratified analysis was completed only if age-by-platform interaction was nominally significant.

## Discussion

In this analysis of survey data, we found that some forms of social media use – in particular, Snapchat, Facebook, and YouTube - were associated with greater levels of self-reported depressive symptoms on a subsequent survey. Notably, with the exception of Snapchat, this association was not explained in regression models by use of other news sources, suggesting it is not accounted for by differential media consumption more broadly. Likewise, the association was not meaningfully changed by number of social supports or face-to-face social interactions at baseline, suggesting it is not mediated by reduction in social interactions more broadly.

While we cannot directly test causation, as in a prior study of Facebook discontinuation^6^, our design allows us to look at incident self-reported depression *following* social media use. They extend prior observations of elevated depressive symptoms in adolescents^1^ or young adults^2^ associated with social media use in cross-sectional studies. They are also consistent with 2 prior short-term longitudinal studies^4,5^. Beyond the ability to look longitudinally, the present work also extends prior studies by examining a broader age range, with a mean age of 56 years. It suggests that the associations previously observed with depressive symptoms are not limited to young adults; indeed, while effect sizes for Facebook are greater among younger adults, effect sizes for TikTok and Snapchat are greater among those age 35 or older.

## Limitations

Results are limited by the inability to control for all potential confounding, lack of dose-response data, and inability to measure nature of social media use which may moderate its impact^8^. Notably, social media use may simply be a marker of underlying vulnerability to depression. On the other hand, one investigation found that viewing one’s own Facebook profile was associated with increased physiologic stress response^9^. The extent to which these findings generalize beyond the COVID-19 pandemic period also remains to be determined; it may be that the impact of social media use is specific to content viewed, for example. A further caveat is that only a (nonrandom) subset of participants return for a second survey, which could yield a biased sample with less generalizability. As the surveys are not labeled as COVID-19 specific, however, the likelihood is lower that these results reflect a selection bias in which individuals with greater interest in, or impact of, COVID-19 preferentially respond. Nonetheless, our findings complement those using cross-sectional designs^2^ or small longitudinal cohorts^4,5^ and extend them to older adults, indicating the need for further investigation of the relationship between social media use and mental health.

## Conclusion

In this survey study of US adults, we identified associations between type of social media utilization at initial survey, and greater levels of depressive symptoms on a subsequent survey.

## Data Availability

Data are not available for distribution.

## Acknowledgements

This study was supported by the National Science Foundation (SES-2029292 and SES-2029792; Drs. Baum, Lazer, Druckman, and Ognyanova). Dr. Perlis is supported by the National Institute of Mental Health (R01MH116270; Dr. Perlis). The sponsors did not contribute to design and conduct of the study; collection, management, analysis, and interpretation of the data; preparation, review, or approval of the manuscript; and decision to submit the manuscript for publication. Dr. Perlis had full access to all the data in the study and takes responsibility for the integrity of the data and the accuracy of the data analysis.

## References

1. Woods HC, Scott H. #Sleepyteens: Social media use in adolescence is associated with poor sleep quality, anxiety, depression and low self-esteem. J Adolesc. 2016;51:41–49. doi:10.1016/j.adolescence.2016.05.008

2. Lin L yi, Sidani JE, Shensa A, et al. Association between Social Media Use and Depression among U.S. Young Adults. Depress Anxiety. 2016;33(4):323–331. doi:10.1002/da.22466

3. Parry DA, Davidson BI, Sewall CJR, Fisher JT, Mieczkowski H, Quintana DS. A systematic review and meta-analysis of discrepancies between logged and self-reported digital media use. Nat Hum Behav. Published online May 17, 2021:1-13. doi:10.1038/s41562-021-01117-5

4. Kross E, Verduyn P, Demiralp E, et al. Facebook Use Predicts Declines in Subjective Well-Being in Young Adults. PLOS ONE. 2013;8(8):e69841. doi:10.1371/journal.pone.0069841

5. Sagioglou C, Greitemeyer T. Facebook’s emotional consequences: Why Facebook causes a decrease in mood and why people still use it. Comput Hum Behav. 2014;35:359–363. doi:10.1016/j.chb.2014.03.003

6. Allcott H, Braghieri L, Eichmeyer S, Gentzkow M. The Welfare Effects of Social Media. Am Econ Rev. 2020;110(3):629–676. doi:10.1257/aer.20190658

7. Kroenke K. Enhancing the clinical utility of depression screening. CMAJ Can Med Assoc J. 2012;184(3):281–282. doi:10.1503/cmaj.112004

8. Burke M, Kraut RE. The Relationship Between Facebook Use and Well-Being Depends on Communication Type and Tie Strength. J Comput-Mediat Commun. 2016;21(4):265–281. doi:10.1111/jcc4.12162

9. Cipresso P, Mauri M, Semonella M, et al. Looking at One’s Self Through Facebook Increases Mental Stress: A Computational Psychometric Analysis by Using Eye-Tracking and Psychophysiology. Cyberpsychology Behav Soc Netw. 2019;22(5):307–314. doi:10.1089/cyber.2018.0602

